# Non-coding variants upstream of *MEF2C* cause severe developmental disorder through three distinct loss-of-function mechanisms

**DOI:** 10.1101/2020.11.15.20229807

**Authors:** Caroline F Wright, Nicholas M Quaife, Laura Ramos-Hernández, Petr Danecek, Matteo P Ferla, Kaitlin E Samocha, Joanna Kaplanis, Eugene J Gardner, Ruth Y Eberhardt, Katherine R Chao, Konrad J Karczewski, Joannella Morales, Meena Balasubramanian, Siddharth Banka, Lianne Gompertz, Bronwyn Kerr, Amelia Kirby, Sally A Lynch, Jenny EV Morton, Hailey Pinz, Francis H Sansbury, Helen Stewart, Britton D Zuccarelli, Genomics England Research Consortium, Stuart A Cook, Jenny C Taylor, Jane Juusola, Kyle Retterer, Helen V Firth, Matthew E Hurles, Enrique Lara-Pezzi, Paul JR Barton, Nicola Whiffin

## Abstract

Clinical genetic testing of protein-coding regions identifies a likely causative variant in only ∼35% of severe developmental disorder (DD) cases. We screened 9,858 patients from the Deciphering Developmental Disorders (DDD) study for *de novo* mutations in the 5’untranslated regions (5’UTRs) of dominant haploinsufficient DD genes. We identify four single nucleotide variants and two copy number variants upstream of *MEF2C* that cause DD through three distinct loss-of-function mechanisms, disrupting transcription, translation, and/or protein function. These non-coding variants represent 23% of disease-causing variants identified in *MEF2C* in the DDD cohort. Our analyses show that non-coding variants upstream of known disease-causing genes are an important cause of severe disease and demonstrate that analysing 5’UTRs can increase diagnostic yield, even using existing exome sequencing datasets. We also show how non-coding variants can help inform both the disease-causing mechanism underlying protein-coding variants, and dosage tolerance of the gene.

## Introduction

The importance of non-coding regulatory variation in common diseases and traits has long been appreciated, however, the contribution of non-coding variation to rare disease remains poorly understood^1–4^. Consequently, current clinical testing approaches for rare disease focus almost exclusively on regions of the genome that code directly for protein, within which we are able to relatively accurately estimate the effect of any individual variant. Using this approach, however, we currently only identify a likely disease-causing variant in around 35% of individuals with severe developmental disorders (DD)^5^. In previous work, we assessed the role of *de novo* mutations (DNMs) in distal regulatory elements, and estimated that 1-3% of undiagnosed DD cases carry pathogenic DNMs in these regions^1^.

Untranslated regions (UTRs) at the 5’ and 3’ end of genes present a unique opportunity to expand genetic testing outside of protein coding regions given they have important regulatory roles in controlling both the amount and location of mRNA in the cell, and the rate at which it is translated into protein^6,7^. Crucially, we also know the genes/proteins that these regions regulate. Given that UTRs account for around the same genomic footprint as protein-coding exons, they have substantial potential to harbour novel Mendelian diagnoses^8,9^.

Recently, we demonstrated that variants creating upstream start codons (uAUGs) in 5’UTRs are under strong negative selection, and are an important cause of Mendelian diseases, including neurofibromatosis and Van der Woude syndrome^10,11^. Initiation of translation at a newly created uAUG can decrease translation of the downstream coding sequence (CDS). The strength of negative selection acting on uAUG-creating variants varies depending on both the match of the sequence surrounding the uAUG to the Kozak consensus, which is known to regulate the likelihood that translation is initiated^12,13^, and the nature of the upstream open reading frame (uORF) that is created. Variants that result in ORFs which overlap the CDS have a larger impact on CDS translation and hence are more deleterious^10,14^.

Here, we screened 9,858 probands from the Deciphering Developmental Disorders (DDD)^5^ study for DNMs in the 5’UTRs of known dominant DD genes. We uncover novel likely disease causing variants that are entirely non-coding, and show how these variants cause disease through three distinct loss-of-function mechanisms. We further analyse the coverage across UTRs in the DDD exome sequencing dataset to demonstrate how these regions can be readily screened in existing datasets to increase diagnostic yield and glean insight into disease causing mechanisms.

## Results

### Identifying de novo 5’UTR variants in DD cases

To investigate the contribution of uAUG-creating variants to severe DD cases, we analysed high-confidence DNMs identified in exome sequencing data from 9,858 parent-offspring trios in the DDD study^5^. Although the vast majority of DNMs identified are coding, as expected with exome sequencing data, some non-coding variants are detectable, particularly near exon boundaries. Given that uAUG-creating variants that decrease CDS translation would only be expected to be deleterious in genes that are dosage sensitive, we restricted our analysis to the 5’UTRs of 359 known haploinsufficient developmental disorder genes from the curated DDG2P database^15^ (Supplementary Table 1).

**Table 1:** Details of *MEF2C* uAUG-creating and upstream deletion variants discussed in this work. Shown are the four uAUG SNVs identified in DDD, uAUG SNVs observed in gnomAD v3.0, and non-coding CNVs found upstream of *MEF2C* in DDD. oORF = overlapping ORF; uORF = upstream ORF; AC = allele count.

| variant (GRCh37) | cDNA description (ENST00000504921.7) | variant effect | kozak strength | patient count | gnomAD v3.0 AC |
| --- | --- | --- | --- | --- | --- |
| *uUAG-creating de novo variants discovered in probands with DD:* |  |  |  |  |  |
| chr5:88119671 T>A | -66A>T | out-of-frame oORF created | moderate | 1 | - |
| chr5:88119708 C>T | -103G>A | out-of-frame oORF created | weak | 1 | - |
| chr5:88119613 G>A | -8C>T | CDS-elongating | strong | 3 | - |
| chr5:88119631 G>A | -26C>T | CDS-elongating | moderate | 3 | - |
| *uAUG-creating variant present in gnomAD:* |  |  |  |  |  |
| chr5:88178869 G>A | -240C>T | uORF created | weak | 0 | 1 |
| chr5:88178876 G>A | -247C>T | uORF created | weak | 0 | 6 |
| *Upstream de novo CNVs identified in probands with DD:* |  |  |  |  |  |
| chr5:88133089-88427361 del | - | promoter and partial 5'UTR deletion | - | 1 | - |
| chr5:88123099-88220350 del | - | promoter and partial 5'UTR deletion | - | 1 | - |

We identified five unique uAUG-creating *de novo* single nucleotide variants (SNVs) in five unrelated probands upstream of two different genes. All of these variants are absent from the Genome Aggregation Database (gnomAD) population reference dataset (both v2.1.1 and v3.0)^16^. Notably, four of the five variants were found in the 5’UTR of *MEF2C* in probands with phenotypes consistent with *MEF2C* haploinsufficiency (Table 1). Two of these DNMs create uAUGs out-of-frame with the *MEF2C* CDS, which are expected to reduce downstream protein translation, whilst the other two create uAUGs in-frame with the CDS, which are expected to elongate the protein (Figure 1). The fifth variant was located in a strong Kozak consensus upstream of *STXBP1* (ENST00000373302.8:c.-26C>G), creating an uAUG out-of-frame with the *STXBP1* CDS; the phenotype of the proband with this variant is consistent with *STXBP1* haploinsufficiency^17^, including global developmental delay, microcephaly, and delayed speech and language development.

**Figure 1:**
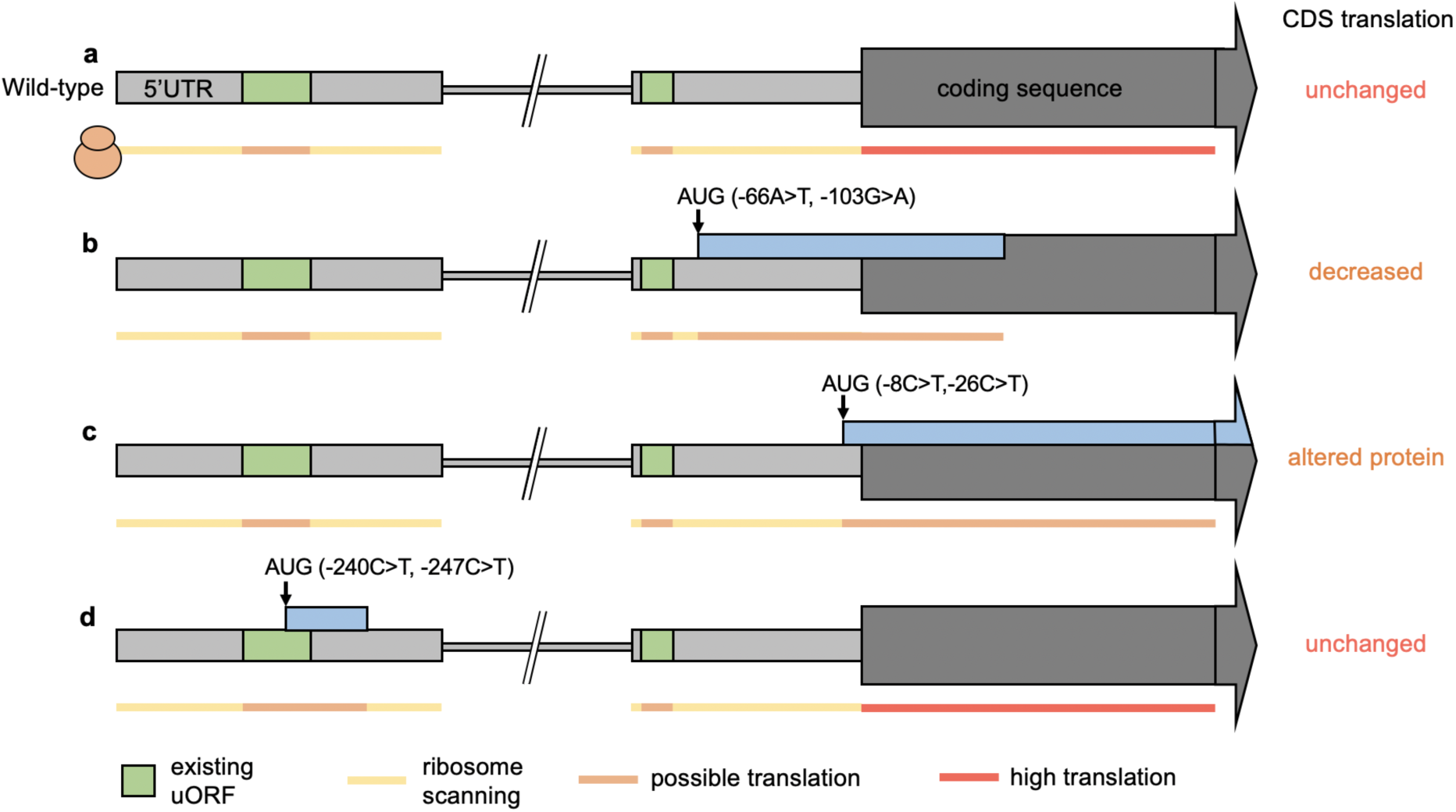
Schematic of the wild-type *MEF2C* gene (a) and the position and effect of uAUG-creating variants identified as *de novo* in developmental disorder cases (b and c) and in gnomAD population controls (d). Upstream open reading frames (uORFs) already present in the sequence are shown in green. Variant positions are represented by arrows. (b) Two case variants create novel ORFs that overlap the coding sequence (CDS) out-of-frame (oORF-creating). If translation initiates at the uAUG, the ribosome will not translate the CDS. (c) Two recurrent case variants create uAUGs in-frame with the CDS. If translation initiates at this uAUG, an elongated protein will be translated. (d) Two variants identified in gnomAD create uORFs far upstream of the CDS which would not be predicted to disrupt translation of the normal protein.

Given the identification of multiple uAUG-creating *de novo* SNVs in *MEF2C* in the DDD study, we subsequently queried high-confidence DNMs identified in 18,789 trios with DD that were exome sequenced by GeneDx^5^ for additional *MEF2C* DNMs. We uncovered three additional *de novo* occurrences of two of the uAUG-creating variants observed in the DDD study. In addition, we identified a further *de novo* occurrence of one of these variants in a DD proband in the UK 100,000 Genomes Project^18^(Table 1).

We also found two additional probands with *de novo* copy number variants (CNVs) overlapping the 5’UTR of *MEF2C* in the DDD study (Table 1). These non-coding variants delete the first exon of the *MEF2C* 5’UTR and >40kb of immediately upstream sequence (248kb and 41kb, respectively), removing the entire promoter (as defined by the Ensembl regulatory build^19^ and H3K4me3 peaks from ENCODE^20^) and likely abolishing transcription of this allele (Supplementary Figure 1). There are no large deletions (>600bps) in this upstream region in the gnomAD structural variant dataset (v2.1)^21^. Non-coding deletions further upstream of *MEF2C* that are predicted to disrupt enhancer function have been identified in DD patients previously^22^.

### De novo 5’UTR variants cause phenotypes consistent with MEF2C haploinsufficiency

We collated all available clinical data for the ten probands with *MEF2C* 5’UTR *de novo* variants and in each case the observed phenotype is consistent with previously reported *MEF2C* haploinsufficiency^23,24^. Specifically, of the nine patients for which detailed phenotypic information was available, the following features were noted: global developmental delay (9/9) with delayed or absent speech (9/9), seizures (8/9), hypotonia (5/9) and stereotypies (2/9). These patients had no other likely disease-causing variants in the coding sequence of *MEF2C*, or in any other known DD genes following exome sequencing.

### uAUG-creating SNVs cause loss-of-function by reducing translation or disrupting protein function

The four uAUG-creating SNVs identified in *MEF2C* result in two different downstream effects. We set about validating the impact of each variant through two distinct experimental approaches: plasmid vectors to evaluate translation of the MEF2C protein, and expression constructs to evaluate MEF2C-dependent transactivation of other genes.

Two of the variants (−66A>T and -103G>A), each found in a single proband, create uAUGs that are out-of-frame with the coding sequence (CDS), creating an overlapping ORF (oORF) that terminates 128 bases after the canonical start site (Figure 1b). Using plasmid vectors with wild-type or mutant 5’UTR sequence cloned upstream of a luciferase reporter gene, we show that both variants result in a significant decrease in translational efficiency (Figure 2a; Supplementary Figure 2a). The amount by which translation is reduced appears to be dependent on the uAUG match to the Kozak consensus sequence, consistent with previous observations^10^. The -103G>A variant, which creates an uAUG with a weak Kozak consensus, results in only a moderate decrease in luciferase expression, and the proband with this variant displays a milder phenotype on clinical review. To validate that this difference in effect is indeed due to the differing Kozak strengths, we mutated a single base in the -103G>A vector to alter the surrounding context to a moderate Kozak consensus match (see online methods). This modification resulted in significantly decreased translational efficiency compared to the unmodified -103G>A variant, to a level equivalent to the -66A>T variant (Supplementary Figure 3). The patient carrying the -103G>A does not have any other 5’UTR variants that could similarly modify the variant’s effect. These data suggest that MEF2C is sensitive to even partial loss-of-function.

**Figure 2:**
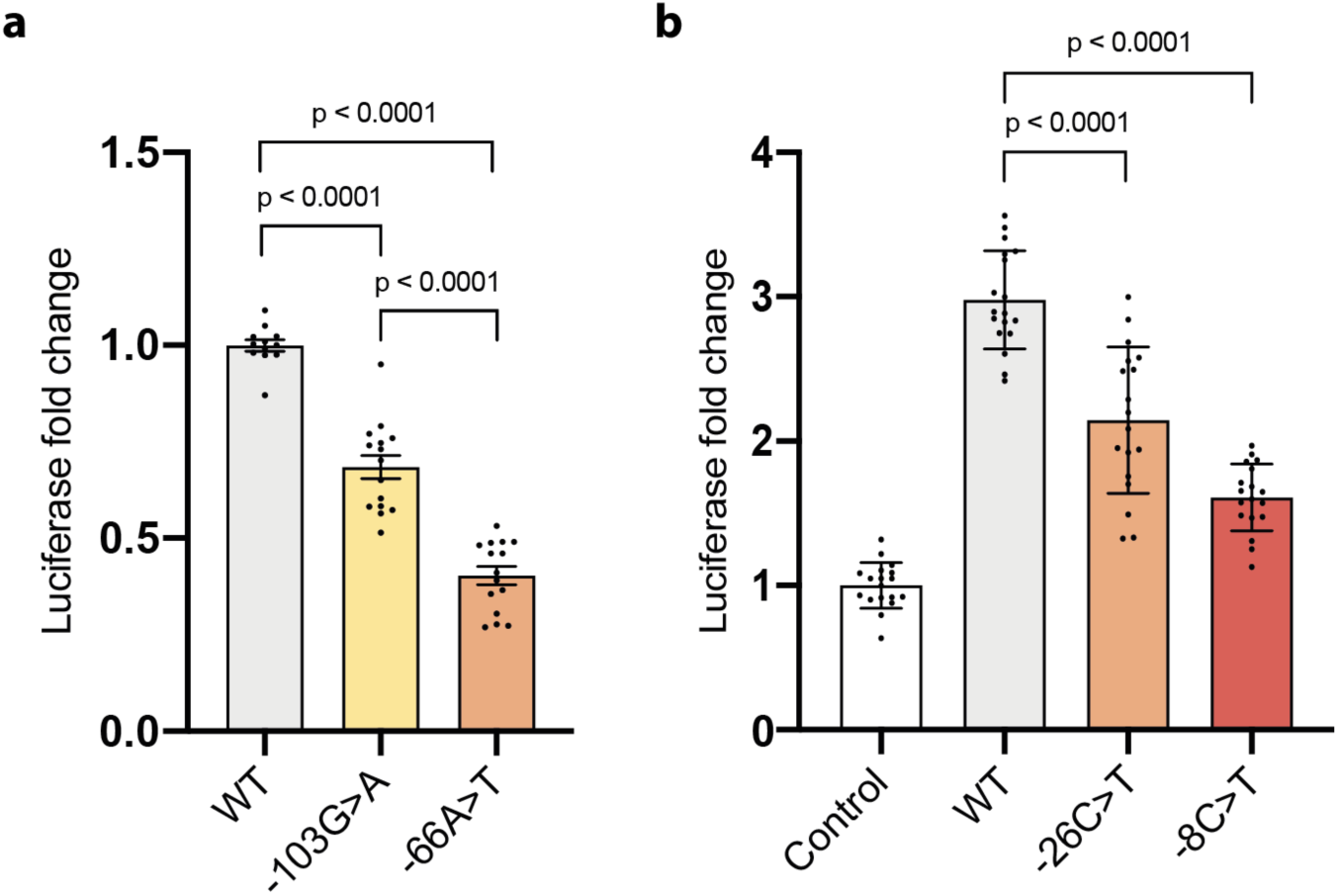
uAUG-creating variants decrease translation of MEF2C (a) or transactivation of target genes (b). (a) *MEF2C* 5’ UTR out-of-frame overlapping ORF (oORF)-creating variants -103G>A and - 66A>T (Figure 1b) reduce downstream luciferase expression relative to wild-type (WT) 5’ UTR in a translation reporter assay. Reduction is stronger for -66A>T (moderate uAUG Kozak context) than for-103G>A (weak Kozak context). (b) Overexpression of MEF2C with the WT 5’ UTR/CDS induces expression of luciferase from a MEF2C-dependent enhancer-luciferase reporter construct, compared to an empty pcDNA3.1 vector control. The *MEF2C* N-terminus-extending variants -26C>T (9AA) and - 8C>T (3AA; Figure 1c) both reduce transactivation. For (a) and (b) bars are coloured by Kozak consensus: yellow=weak; orange=moderate; red=strong.

The other two variants (−8C>T and -26C>T) are both observed recurrently *de novo*, each in three unrelated probands (Table 1). Both variants create uAUGs that are in-frame with the CDS, resulting in N-terminal extensions of three and nine amino acids respectively (Figure 1c). MEF2C is a transcription factor, and critical to its function is the DNA-binding domain located at the extreme N-terminal region^25^. Although no structure is available for the MEF2C protein, numerous crystal and NMR structures of the N-terminal DNA-binding domain of human MEF2A are available, which is 96% identical in sequence to MEF2C. These structures show clearly that the extreme N-terminus of the protein is in direct contact with DNA^26,27^, and that the first few residues bind directly into the minor groove (Figure 3). We assayed MEF2C-dependent transactivation using MEF2C expression constructs with wild-type and mutant 5’UTR sequences. These data demonstrate significantly reduced activation of target gene transcription from the variants (Figure 2b; Supplementary Figure 2b and c), compared to wild-type MEF2C. Once again, the strength of the effect is dependent on the uAUG context, with the -8C>T variant that creates a strong Kozak consensus having a larger effect, almost abolishing transactivation activity.

**Figure 3:**
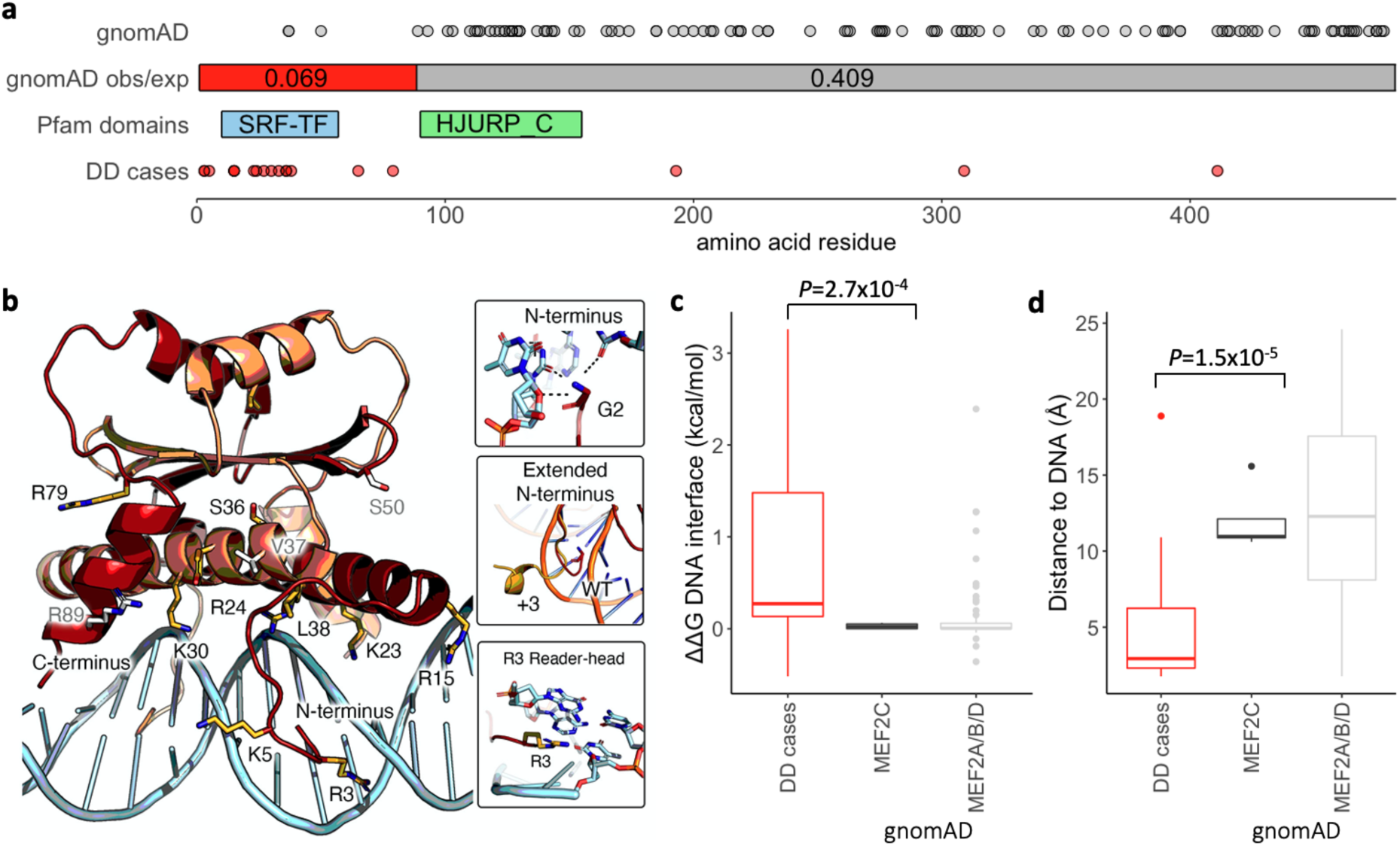
(a) The N-terminal region of *MEF2C* is highly-constrained for missense variants in gnomAD (obs/exp=0.069), with much lower constraint across the rest of the protein (obs/exp=0.41). This region of high constraint correlates with the location of the majority of *de novo* missense variants identified in DD cases (red circles), while gnomAD variants are mostly outside of this N-terminal region (grey circles). (b) The N-terminal portion of the MEF2C dimer [1-92], modelled using structures of the human MEF2A dimer which is 96% identical in sequence to MEF2C, bound directly to its consensus DNA sequence. Side chains of amino acids with pathogenic *de novo* missense variants from DDD, GeneDx and ClinVar are shown in yellow, with gnomAD *MEF2C* missense variants in grey. Most pathogenic missense variants either protrude directly into the DNA or are located in the DNA-binding helix. In particular, the terminal amine (Gly2, top inset) along with Arg3 (bottom inset) act as reader-heads for nucleobase specificity, which is likely disrupted in the N-terminal extension variants (middle inset). All pathogenic and gnomAD variants can be viewed in our interactive protein structure browser here: https://michelanglo.sgc.ox.ac.uk/r/mef2c. (c-d) Missense variants from DD cases (DDD, GeneDx and ClinVar) are significantly more disruptive to the interaction with DNA as measured by ΔΔG values (c) and closer to the bound DNA molecule (d) than *MEF2A-D* variants in gnomAD (see online methods).

We looked in the gnomAD dataset^16^ for uAUG-creating variants that might have similar impacts. Across the exome (v2.1.1) and genome (v3.0) sequencing datasets, there are only two uAUG-creating variants in the *MEF2C* 5’UTR. Crucially, neither of these fall into the proximal 5’UTR exon and neither create ORFs overlapping the CDS. In both instances, the uAUGs are created into weak Kozak-consensus contexts, and they have in-frame stop codons after 6bps (allele count = 6) and 57bps (allele count = 1) respectively (Table 1; Figure 1d). These variants would therefore not be expected to have substantial, if any, effect on *MEF2C* translation.

### Pathogenic de novo missense variants likely cause loss-of-function of MEF2C through disrupting DNA-binding

Whilst the major recognised mechanism through which pathogenic variants in *MEF2C* lead to severe developmental phenotypes is loss-of-function, *de novo* missense variants are also significantly enriched in DD trios (*P*=1.3×10^−14^)^5^ and multiple pathogenic missense variants are reported in ClinVar^28^. These variants are almost exclusively found at the extreme N-terminus of the protein (Supplementary Table 2), in the DNA-binding region, which is also highly constrained for missense variants in gnomAD (obs/exp=0.069; calculated on 125,748 exome sequenced samples in v2.1.1; Figure 3a). We hypothesised that these pathogenic missense variants are also causing loss-of-function by disrupting DNA-binding of MEF2C as has been demonstrated for random disruptions to the N-terminal region previously^25^. Using the structure of the N-terminal MEF2A homodimer bound to DNA, we modelled the location of pathogenic missense variants in MEF2C, as well as missense variants in gnomAD v2.1.1 across all members of the myocyte enhancer factor 2 protein family (MEF2A-D; 84% N-terminal domain sequence identity; Supplementary Table 3; Supplementary Figure 4), and saw a significant enrichment of pathogenic variants interacting directly with DNA via both the N-terminal loop and DNA-binding helix (Fisher’s *P*=2.6×10^−5^, Figure 3b; Supplementary Tables 4 & 5). We further calculated the change in Gibbs free energy (ΔΔG) of both the protein-DNA interaction and the complex stability for each missense change. Variants found in DD cases have significantly increased ΔΔG scores compared to gnomAD variants (Wilcoxon *P*=2.7×10^−4^; Figure 3c) and are significantly closer to the bound DNA (Wilcoxon *P*=1.5×10^−5^; Figure 3d; Supplementary Table 6). Together, these data suggest that disease-causing missense variants in *MEF2C* act through a loss-of-function mechanism. Indeed, excluding the N-terminal DNA-binding domain, the remainder of *MEF2C* shows much weaker constraint against missense variants in gnomAD (obs/exp=0.41), and only nominal enrichment for *de novo* missense variants in DD cases (*P*=0.041).

### Disease-causing 5’UTR variants can be detected in exome sequencing data

Given our ability to identify 5’UTR variants in *MEF2C*, we investigated the extent to which these regions are captured across all genes in the exome sequencing dataset from the DDD study. We find that 30.7% of all gene 5’UTR bases and 20.4% of 5’UTR bases of our DD haploinsufficient genes (average of 73 bps per gene; n=345 with MANEv0.91 transcripts) are covered at a mean coverage threshold of >10x. The average length of 5’UTRs in DD haploinsufficient genes is 356 bps (Figure 4a), with 42.0% containing multiple exons (Figure 4b). As expected, 5’UTR coverage decays as distance from the CDS increases (Figure 4c), with distal exons very poorly covered (6.7% of bases >10x). In comparison, a much lower proportion of 3’UTR bases (6.0%) are covered at >10x, which is unsurprising given that 3’UTRs are much longer than 5’UTRs, at an average of 2,652 bps for our DD haploinsufficient genes.

**Figure 4:**
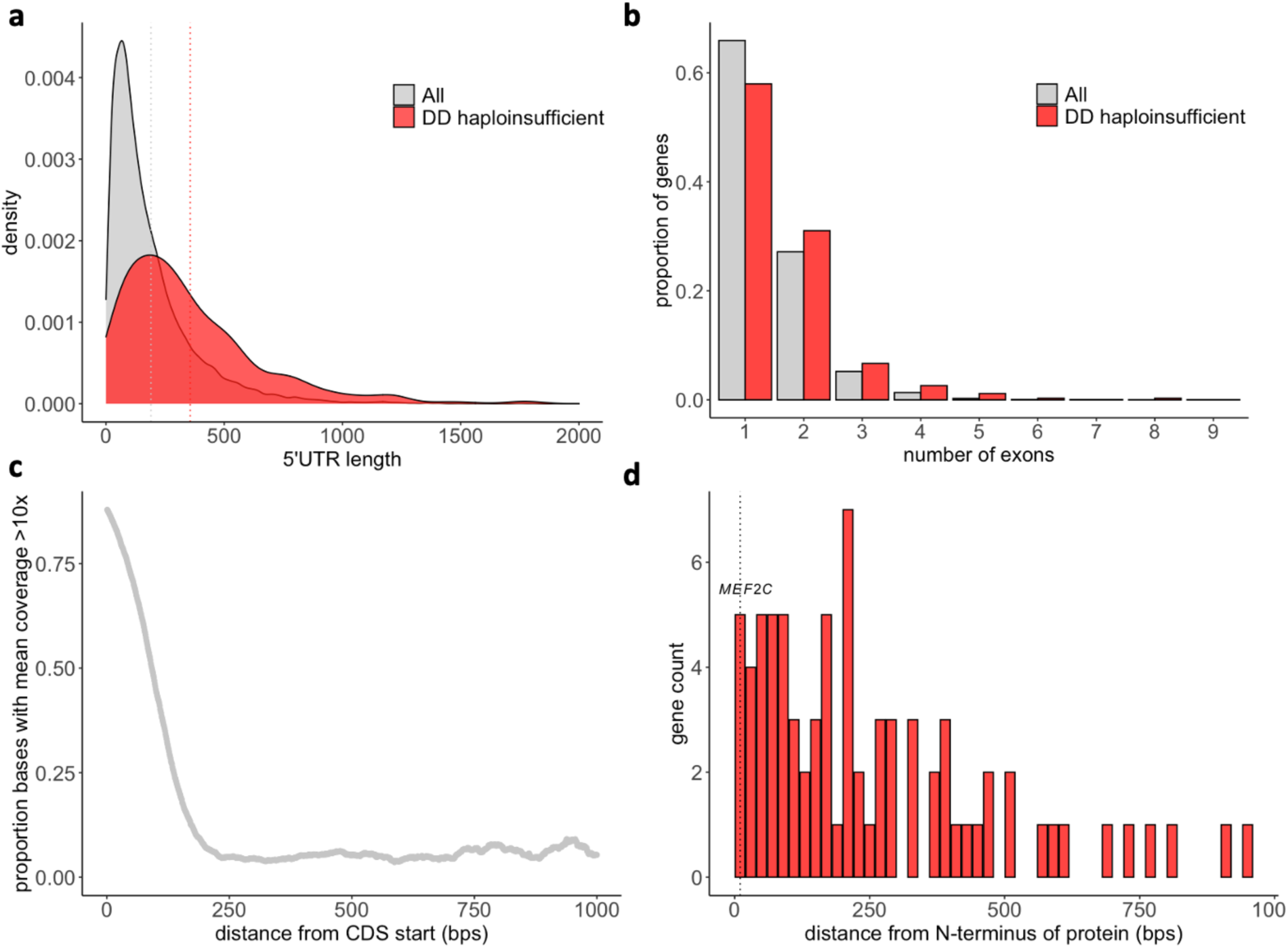
5’UTRs of DD haploinsufficient genes (red) are longer (a), and a higher proportion have multiple exons (b) compared to 5’UTRs of all genes (grey). Mean lengths for each gene set in (a) are shown as dotted lines. (c) The coverage of 5’UTRs decays rapidly with distance from the CDS (x-axis truncated at 1000 bps). (d) The position of DNA-binding domains (including homeodomains, zinc-fingers, and specific DNA-binding domains) in DD haploinsufficient genes with respect to the N-terminus of the protein; MEF2C is one of three proteins with a DNA-binding domain that starts within 10 bps of the N-terminus.

We identified 3,962 possible uAUG-creating variants in DD haploinsufficient genes that would create out-of-frame overlapping ORFs (n=2,782) or CDS-elongations (n=1,180). Of these, 42.4% are sequenced at >10x coverage across the DDD study dataset (40.2% of out-of-frame and 47.6% of CDS-elongating). However, we would not expect CDS-elongating variants to cause a loss-of-function for the majority of genes. Rather, we expect this to be limited to genes with important functional domains at the extreme N-terminus that would be adversely affected by the addition of extra N-terminal amino acids, either through disrupting binding or altering protein structure. Based on Pfam domain predictions, only three of the proteins encoded by our 359 DD haploinsufficient genes, including MEF2C, have DNA-binding domains that start within 10 bps of the N-terminus (Figure 4d); the other two (*ZNF750* and *SIM1*) encode an N-terminal zinc-finger and basic helix-loop-helix, respectively, and although no structures are available, these bind DNA via specific motifs that are unlikely to include the extreme N-terminal residues.

## Discussion

Here, we have identified six non-coding DNMs in *MEF2C* in ten patients with severe developmental disorders. These variants act via three distinct loss-of-function mechanisms at different stages of expression regulation: (1) two large deletions remove the promoter and part of the 5’UTR and are predicted to abolish normal transcription of *MEF2C*; (2) two SNVs create out-of-frame uAUGs and reduce normal translation of the *MEF2C* coding sequence; and (3) two SNVs create in-frame uAUGs that elongate the *MEF2C* coding sequence, disrupting binding of the MEF2C protein to DNA and reducing subsequent transactivation of gene-expression.

These observations demonstrate the importance of screening 5’UTRs of known disease genes in patients that remain genetically undiagnosed. We have previously identified 20 DNMs (15 SNVs and 5 CNVs) impacting *MEF2C* protein-coding regions in the 9,858 DDD family trios analysed. The six additional non-coding DNMs described here therefore comprise 23% of known diagnostic variants impacting *MEF2C* in this cohort.

Our data show that 5’UTR variants can be identified in existing datasets that were primarily designed to capture coding sequences, with 30.7% of 5’UTR bases having sufficient (>10x) coverage in exome sequencing data from the DDD study. However, exome sequencing data is likely to only identify UTR variants that are proximal to the first and last exons of genes, and whole genome or expanded panel sequencing will be required to assay distal or poorly covered UTRs. Furthermore, given their large size, 3’UTRs are particularly poorly covered in exome sequencing datasets. There are examples of disease-causing variants within 3’UTRs, including those impacting polyA signals and microRNA binding^8,29–31^, which will not be detected using these methodologies but that could increase diagnostic yield.

Although we screened DNMs in the 5’UTRs of a set of 359 known haploinsufficient DD genes, four of the five identified *de novo* uAUG-creating variants were found in *MEF2C*. This enrichment in a single gene is likely due to a combination of factors (Supplementary Figure 5). Firstly, *MEF2C* has a proximal 5’UTR exon that is very well covered in the DDD exome sequencing data. Secondly, this 5’UTR exon contains a large number of sites where a variant could create an uAUG, with only two DD haploinsufficient genes having more well-covered possible uAUG-creating sites. Thirdly, unlike the other genes with well-covered possible uAUG-creating sites, *MEF2C* haploinsufficiency is a recurrent cause of DD within the DDD study (Supplementary Figure 5). Finally, due to the direct interaction of the extreme N-terminus of MEF2C with DNA, CDS-elongating variants are also likely to be pathogenic, which is unlikely to be the case in the vast majority of other haploinsufficient DD genes. As a result, *MEF2C* may be unusual in its potential for pathogenic mutations in the 5’UTR and similarly large increases in diagnostic yield are unlikely across most DD haploinsufficient genes. Nethertheless the enrichment of uAUG-creating variants in *MEF2C* is striking: only 14 of 426 possible variants create uAUGs (at 142 5’UTR bases that are well-covered in the DDD study exome sequencing data), yet all four DNMs observed in the DDD study in the *MEF2C* 5’UTR are uAUG-creating (binomial *P*=1.2×10^−6^).

As we extend our analyses to detect non-coding variants, we caution that interpretation of UTR variants still remains a critical challenge. Every 5’UTR has a unique combination of regulatory elements tightly regulating RNA stability and protein expression^32,33^, and the impact of any variant will vary with the gene-specific context. Functional validation of identified variants will therefore be crucial to prove (or reject) causality. Some variants may have only a partial regulatory effect, but these variants can nonetheless be harnessed to assess the extent to which perturbation of protein levels or function is tolerated, potentially leading to reduced expressivity and/or lower penetrance. In the case of *MEF2C*, our results suggest that even partial reductions in protein expression lead to severe disease.

Finally, we note how the mechanism of action of non-coding variants can inform the mechanisms underlying protein-coding variants. Identification and characterisation of the effect of the CDS-elongating *MEF2C* variants led us to analyse the domain structure of MEF2C protein and confirm that all the currently identified missense variants likely also act via disrupting DNA-binding, leading to a loss-of-function. Missense variants identified in other regions of the protein are therefore unlikely to cause severe DD.

In conclusion, our results further highlight the important contribution of non-coding regulatory variants to rare disease and underscore the huge promise of large whole-genome sequencing datasets to both find new diagnoses and further our understanding of regulatory disease mechanisms.

## Data Availability

The DDD study data are available under managed access from the European Genome-phenome Archive (Study ID EGAS00001000775), and likely diagnostic variants are available open access in DECIPHER. Code used for modelling case and population variants on the MEF2C protein structure can be found here: https://github.com/matteoferla/MEF2C_analysis

## Acknowledgements

NW is currently supported by a Sir Henry Dale Fellowship jointly funded by the Wellcome Trust and the Royal Society (Grant Number 220134/Z/20/Z). Initial work was completed whilst NW was supported by a Rosetrees and Stoneygate Imperial College Research Fellowship. NQ is supported by the Imperial College Academic Health Science Centre. This work is additionally supported by The Rosetrees Trust (Grant Number H5R01320), Fondation Leducq [16 CVD 03], the National Institute for Health Research (NIHR) Imperial College Biomedical Research Centre, the Cardiovascular Research Centre, Royal Brompton & Harefield NHS Trust, and the NIHR Oxford Biomedical Research Centre Programme. The views expressed are those of the author(s) and not necessarily those of the NHS, the NIHR or the Department of Health. The DDD study presents independent research commissioned by the Health Innovation Challenge Fund (Grant Number HICF-1009-003) a parallel funding partnership between the Wellcome Trust and the Department of Health, and the Wellcome Trust Sanger Institute (Grant Number WT098051). See Nature 2015;519:223-8 or www.ddduk.org/access.html for full acknowledgement. This research was made possible through access to the data and findings generated by the 100,000 Genomes Project. The 100,000 Genomes Project is managed by Genomics England Limited (a wholly owned company of the Department of Health and Social Care). The 100,000 Genomes Project is funded by the National Institute for Health Research and NHS England. The Wellcome Trust, Cancer Research UK and the Medical Research Council have also funded research infrastructure. The 100,000 Genomes Project uses data provided by patients and collected by the National Health Service as part of their care and support.

## Online methods

### Patient recruitment, sample collection and clinical data

The DDD Study has UK Research Ethics Committee approval (10/H0305/83, granted by the Cambridge South REC, and GEN/284/12 granted by the Republic of Ireland REC). Patients with severe, undiagnosed developmental disorders and their parents were recruited and systematically phenotyped by the 24 Regional Genetics Services within the United Kingdom (UK) National Health Service and the Republic of Ireland. Saliva samples were collected from probands and parents, and DNA extracted as previously described^34^; blood-extracted DNA was also collected for probands where available. Clinical data (growth measurements, family history, developmental milestones, etc.) were collected using a standard restricted-term questionnaire within DECIPHER^35^. Detailed phenotype data for the patients in this study is available on request.

### Genetic data

Array-CGH analysis was performed using 2 x 1M probe custom designed microarrays (Agilent; Amadid No.s 031220/031221) as described previously^34^. Exome sequencing was performed using Illumina HiSeq (75-base paired-end sequencing) with SureSelect baits (Agilent Human All-Exon V3 Plus and V5 Plus with custom ELID C0338371) and variants were called and annotated as described previously^34^. We used DeNovoGear^36^ (version 0.54) to detect likely DNMs from trio exome BAM files and Ensembl Variant Effect Predictor^37^ was used to annotate predicted consequences. The data are available under managed access from the European Genome-phenome Archive (Study ID EGAS00001000775), and likely diagnostic variants are available open access in DECIPHER.

### Defining a gene-set of interest

We limited our analysis to 359 DDG2P^15^ genes with a confirmed or probable role in developmental disorders and with a dominant (including X-linked dominant) loss-of-function disease mechanism (downloaded from https://www.ebi.ac.uk/gene2phenotype/downloads on 21st July 2020; Supplementary Table 1).

### Identifying uAUG-creating variants in DDD

We defined high-confidence DNMs in DD as previously^38^, using the following criteria: minor allele frequency < 0.01 in our cohort and reference databases, depth in the child > 7, depth in both parents > 5, Fisher strand bias p-value > 10^−3^, and a posterior probability of being a DNM from DeNovoGear > 0.00781^36^. Additionally, we filtered out DNMs with some evidence of an alternative allele in one of the parents and indels with a low variant allele fraction (<30% of the reads support the alternative) that had a minor allele frequency > 0. We cross-referenced this list of high-confidence DNMs with a list of all possible uAUG-creating SNVs from previous work^10^. We also assessed any small insertions and deletions that could form uAUGs.

The strength of the Kozak consensus surrounding each uAUG was assessed as described previously^10^. Specifically, we assessed the positions at −3 and +3 relative to the A of the AUG, requiring both the −3 base to be either A or G and the +3 to be G for an annotation of ‘Strong’. if only one of these conditions was true, the strength was deemed to be ‘Moderate’ and if neither was the case ‘Weak’.

### Defining the 5’UTR of MEF2C

We used the MANE select transcript ENST00000504921.7 for which the 5’UTR was defined using CAGE data from the FANTOM5 project^39^ and exon level expression from the GTEx project^40^ (https://www.ncbi.nlm.nih.gov/refseq/MANE/). The 5’UTR of *MEF2C* was therefore defined as two exons: chr5:88178772-88179001 and chr5:88119606-88119747 on GRCh37, or chr5:88882955-88883184 and chr5:88823789-88823930 on GRCh38.

### Searching for MEF2C 5’UTR variants in external datasets

We queried the regions corresponding to the *MEF2C* 5’UTR for *de novo* variants in (1) a set of 18,789 DD trios sequenced by the genetic testing company GeneDx^5^, (2) 13,949 rare disease trios from the main programme v9 release of the Genomics England 100,000 Genomes Project^18^ (https://cnfl.extge.co.uk/display/GERE/De+novo+variant+research+dataset), and (3) variants in the v3.0 dataset of the Genome Aggregation Database (gnomAD)^16^.

### Assessing 5’UTR coverage

Regions corresponding to 5’UTRs were extracted from the .gff file from the MANE project v0.91 (ftp://ftp.ncbi.nlm.nih.gov/refseq/MANE/MANE_human/release_0.91/ ; MANE select transcripts). For each base, we calculated the mean coverage across 1,000 randomly selected samples from DDD. A mean coverage of >10x was used to call a base ‘covered’. Analysis was limited to genes with a defined MANE select transcript. For our DD haploinsufficient genes this was 345/359 genes (96.1%).

To identify all possible uAUG-creating variants in DD haploinsufficient genes, we extracted the 5’UTR sequence from the MANE rna.fna file and used the UTRannotator^41^ to find all possible uAUG-creating sites and annotate their consequence.

### Functional validation of variants creating out-of-frame oORFs: by MEF2C 5’UTR-luciferase assay

#### Expression constructs

WT and variant MEF2C 5’UTRs were cloned directly upstream of GLuc in the pEZX-GA02 backbone (Labomics) and sequenced to confirm integrity. Secreted alkaline phosphatase (SEAP) was expressed on the same construct for normalisation of transfection efficiency.

#### Cull culture, transfection and analysis

HEK293T cells were purchased from ATCC and cultured in Dulbecco’s Modified Eagle Medium (glutamine+, pyruvate+) supplemented with 10% foetal bovine serum and 1% penicillin/streptomycin. Cells were transfected with MEF2C 5’UTR-luciferase constructs using Lipofectamine 3000, following manufacturer’s protocols. After 24h, GLuc and SEAP were simultaneously quantified using the Secrete-Pair Dual Luminescence assay (Genecopoeia).

#### Qpcr

RNA was purified from cells using phenol-chloroform extraction and the Qiagen RNeasy Miniprep kit. RNA quantity was normalised and cDNA generated using IV VILO reverse transcriptase following manufacturer’s protocols. Quantitative PCR was performed using SYBR green master mix on a Quantstudio 7 Real-time PCR system and results normalised to co-amplified GAPDH. The following primers were used: GLUC F: 5’ CTGTCTGATCTGCCTGTCCC 3’, GLUC R: 5’ GGACTCTTTGTCGCCTTCGT 3’, SEAP F: 5’ ACCTTCATAGCGCACGTCAT 3’ and SEAP R: 5’ TCTAGAGTAACCCGGGTGCG 3’, GAPDH F: 5’ GGAGTCAACGGATTTGGTCG 3’, GAPDH R: ATCGCCCCACTTGATTTTGG 3’.

#### Kozak mutagenesis

The kozak context of the -103G>A MEF2C 5’UTR-luciferase vector was modified using the Quikchange II mutagenesis kit, following manufacturers protocols. The following PAGE-purified mutagenesis primers were used: F: 5’CTCCTTCTTCAGCATTTTCACAGCTCAGTTCCCAA 3’, R: 5’ TTGGGAACTGAGCTGTGAAAATGCTGAAGAAGGAG 3’. Constructs were fully sequenced to verify mutation and construct integrity in each case

### Functional validation of CDS-elongating variants: by MEF2 binding site-luciferase transactivation assay

#### Expression and reporter constructs

WT and variant MEF2C 5’ UTR+CDS oligos were cloned into the pReceiver-M02 expression construct (Labomics) and sequenced to confirm integrity. For normalisation of transfection efficiency, cells were co-transfected with pRL-Renilla. A desMEF2-luciferase reporter construct was used to quantify the transactivational efficiency of each MEF2C expression construct, and consisted of three copies of a high-affinity MEF2 binding site^42^, linked to an hsp68 minimal promoter in pGL3 (Promega)^43^.

#### Cell culture and transfection

HL1 cardiomyocytes were cultured in Claycomb medium, supplemented with 2 mM L-glutamine, 10% FBS and 100 g/ml Penicillin/Streptomycin. Culture surfaces were pre-treated with gelatin/fibronectin. Cells were co-transfected with 1) desMEF2-luciferase reporter construct, 2) pRL-Renilla transfection control, and 3) expression construct of either: i) empty pcDNA3.1 (negative control), ii) WT MEF2C 5’ UTR+CDS, iii) MEF2C -26C>T, or iv) MEF2C -8C>T. Transfection was with Lipofectamine 2000, following manufacturers protocols. 48h after transfection, firefly and Renilla Luciferases were quantified by the Promega Dual-Luciferase Reporter Assay System.

#### Western blot

HL1 cells were lysed in RIPA buffer in the presence of protease and phosphatase inhibitors (04693159001 and 04906845001, Roche Diagnostics). Lysates were separated on SDS-PAGE gels and transferred to PVDF membranes, which were blocked with 3% skimmed milk in TBS. The primary antibody was anti-MEF2C (ab211493, Abcam), and the secondary antibody was anti-mouse P0447 from Dako. The membrane was developed using ECL reagent (AC2204, Azure Biosystems) and intensity of the bands quantified using ImageJ software.

#### Statistical analysis for all assays

Data were analysed for statistical significance using 1-way ANOVA followed by Tukey’s post-test, using GraphPad Prism 8.0.

### CNV calling

Four CNV detection algorithms (XHMM^44^, CONVEX^34^, CLAMMS^45^ and CANOES^46^) were used to ascertain CNVs from exome data, followed by a random forest machine learning approach to integrate and filter the results (manuscript in preparation).

Layered H3K4me3 data (to visualise active promoter regions) was downloaded from the UCSC table browser (https://genome.ucsc.edu/cgi-bin/hgTables) for GN12878 as a representative cell line and plotted alongside the identified CNVs in Supplementary Figure 1.

### Modelling missense disruption to DNA-binding

We collated a set of missense variants identified in *MEF2C* in DD cases comprising all *de novo* variants from trios in DDD and GeneDx published previously^5^, and variants from ClinVar either flagged as being identified as *de novo*, or with functional evidence (Supplementary Table 2).

As a comparator, we used missense variants from gnomAD v2.1.1^16^. Given that there are only three variants in the N-terminal region of *MEF2C* in gnomAD, but the sequence of the N-terminal region is near identical across the four MEF2 proteins (Supplementary Figure 4), we used missense variants from all four genes (*MEF2A-D*; Supplementary Table 3).

Based on structures of the N-terminal MADS-box of *MEF2A* homodimer(1egw, 3kov and 6byy, residues 1-92) bound to its DNA consensus sequence^26^, we categorised residues into one of four categories: (1) in N-terminal random coil and in contact with the DNA (2) in N-terminal alpha-helix pointing towards the DNA; (3) in N-terminal alpha-helix pointing away from the DNA; or (4) distal to the DNA contact surface (Supplementary Table 4). We used a two-sided Fisher’s exact test to assess for an enrichment of variants in contact or pointing towards the DNA helix in DD cases (Supplementary Table 5).

The Swissmodel threaded model of MEF2C based upon PDB:6BYY (89% identity)^47,48^ was energy minimised using Pyrosetta^49^ with 15 FastRelax cycles^50^ against the electron density of PDB:6BYY and 5 unconstrained. The DNA was extended on both ends due to the proximity of R15. Mutations were introduced and the 10 Å neighbourhood was energy minimised. Gibbs free energy was calculated using the Rosetta ref2015 scorefunction^51^.

Gibbs free energy of binding was calculated by pulling away the DNA and repacking sidechains and, in the case of residues in the N-terminal loop, thoroughly energy minimising the backbone of the loop as this is highly flexible when unbound. N-terminal extensions were made using the RemodelMover^52^ with residues 2-5 also remodelled as determined by preliminary test. Closest distance of each residue to the DNA was calculated with the Python PyMOL module. Code used for this analysis can be found at: https://github.com/matteoferla/MEF2C_analysis. This interactive page was made in MichelaNGLo^53^.

All missense variants are annotated with respect to the Ensembl canonical transcript ENST00000340208.5.

### Calculating regional missense constraint and de novo enrichment

We determined regional missense constraint by (1) extracting observed variant counts from the 125,748 samples in gnomAD v2.1.1, (2) calculating the expected variant count per transcript, and (3) applying a likelihood ratio test to search for significant breaks that split a transcript into two or more sections of variable missense constraint.

Observed missense variants were extracted from the gnomAD exomes Hail Table (version 2.1.1) as described previously^16^, using the following criteria:

- Annotated as a missense change in a canonical transcript of a protein-coding gene in Gencode v19 by Variant Effect Predictor (VEP, version 85)
- Median coverage greater than 0 in the gnomAD exomes data
- Passed variant filters
- Adjusted allele count of at least one and an allele frequency less than 0.1% in the gnomAD exomes

To calculate the expected variant count, we extended methods described previously^16^ to compute the proportion of expected missense variation per base. Briefly, we annotated each possible substitution with local sequence context, methylation level (for CpGs), and associated mutation rate from the table computed in Karczewski *et al*.^16^ We aggregated these mutation rates across the transcript and calibrated models based on CpG status and median coverage. To determine the expected variants for a given section of the transcript, we calculated the fraction of overall the mutation rate represented by the section and multiplied it by the aggregated expected variant count for the full transcript.

We defined missense constraint by extending the methods from Samocha *et al*.^54^ We employed a likelihood ratio test to compare the null model (transcript has no regional variability in missense constraint) with the alternative model (transcript has evidence of regional variability in missense constraint). We required a χ^2^ value above a threshold of 10.8 to determine significance for each breakpoint, and in the case of multiple breakpoints, retained the breakpoint with the maximum χ^2^. This approach defined a single breakpoint in the *MEF2C* canonical transcript at chr5:88057138 (GRCh37).

To evaluate the enrichment of DNMs in the transcript when removing the N-terminal section, we determined the probability of a missense mutation in that region and then compared the observed number of DNMs (n=3) with the expected count in 28,641 individuals using a Poisson test. Specifically, we took the probability of a missense mutation (mu_mis) as provided in the gnomAD v2.1 constraint files for *MEF2C* and adjusted it for the fraction of mutability represented in the latter section of the gene (∼79.5%).

## Supplementary Figures

**Supplementary Figure 1:**
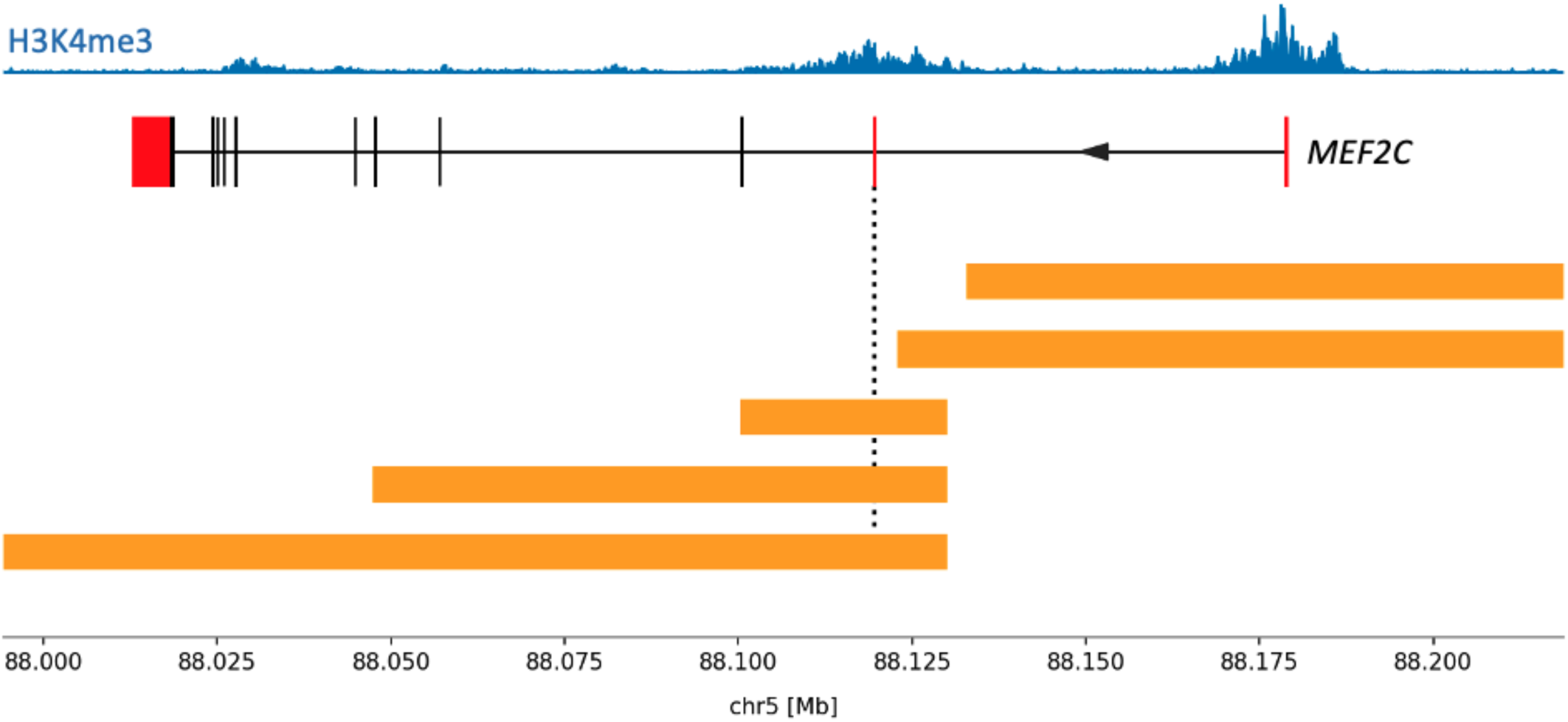
Two non-coding deletions remove the distal 5’UTR exon of *MEF2C* and the entire promoter sequence. Coding exons are shown in black with UTRs in red. The dotted line indicates the start of the coding sequence. The five deletions identified in DDD in the *MEF2C* region are shown as orange bars, the top two of which are entirely non-coding. A representative H3K4me3 dataset from ENCODE is plotted in blue across the top (GN12878) to show active promoter regions.

**Supplementary Figure 2:**
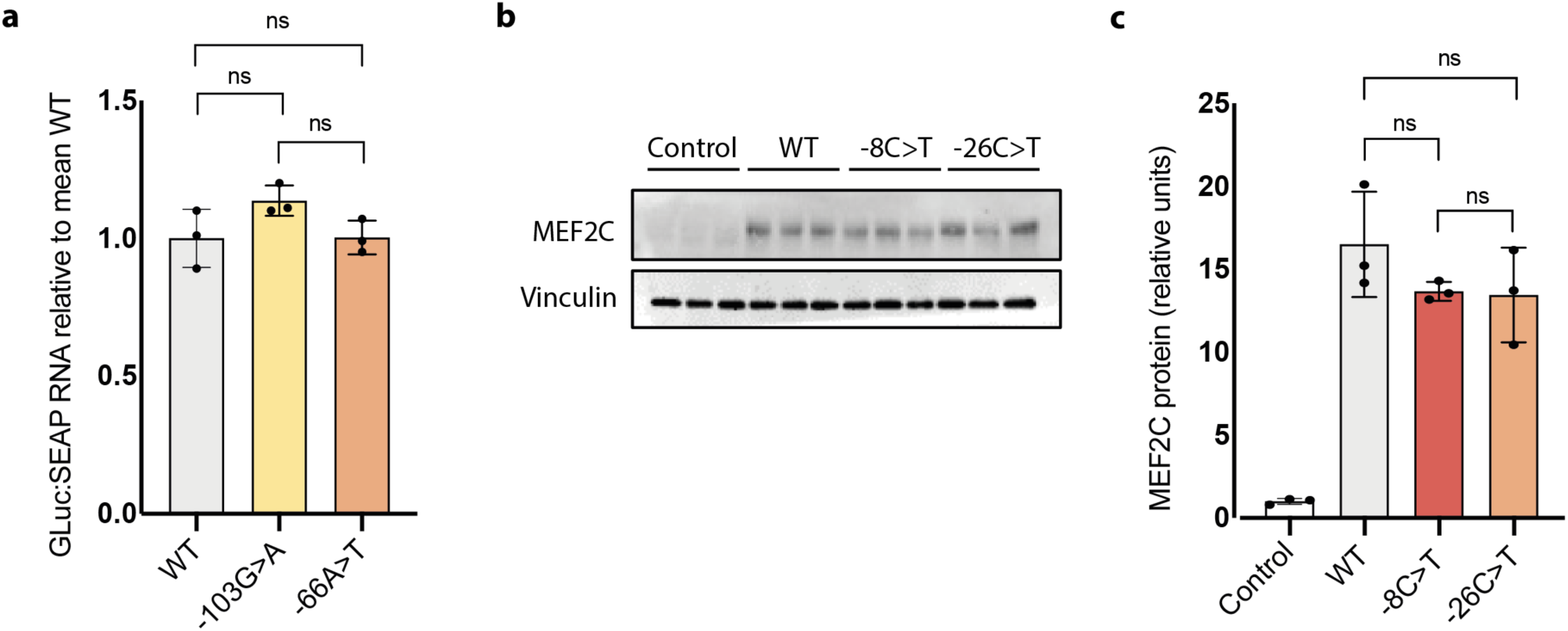
(a) Relative Gaussia luciferase (GLuc) RNA levels remain unchanged with each out-of-frame oORF-creating variant when normalised to RNA of secreted alkaline phosphatase (SEAP) transfection control. (b and c) The decreases in transactivation seen for the CDS-elongating variants -8C>T and -26C>T are not accompanied by a significant change in protein levels. For (a) and (c) bars are coloured by Kozak consensus: yellow = weak; orange = moderate; red = strong. ns = not significant.

**Supplementary Figure 3:**
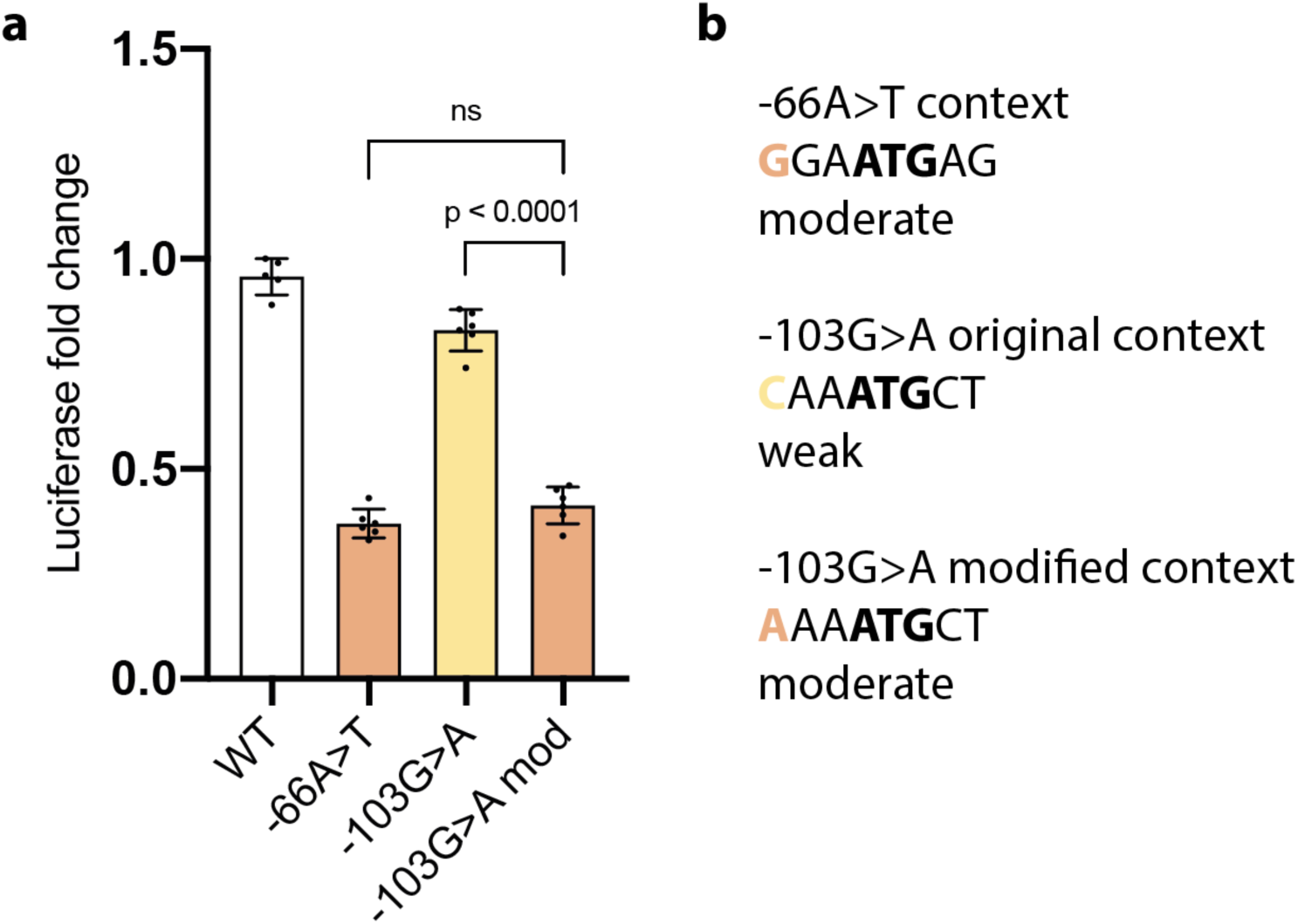
A single base mutation in the context surrounding the -103G>A variant which changes a weak Kozak consensus into a moderate consensus significantly reduces translational efficiency. (a) oORF-creating variants -103G>A and -66A>T reduce downstream luciferase expression relative to wild-type (WT) 5’ UTR in a translation reporter assay. Reduction is stronger for -66A>T (moderate Kozak context) than for -103G>A (weak Kozak context). Modifying the context surrounding the -103G>A variant into a moderate Kozak context (as shown in b) reduces downstream luciferase expression compared to the unmodified vector. The translational efficiency of the modified vector is equivalent to the -66A>T variant which also has a moderate Kozak consensus. ns = not significant.

**Supplementary Figure 4:**
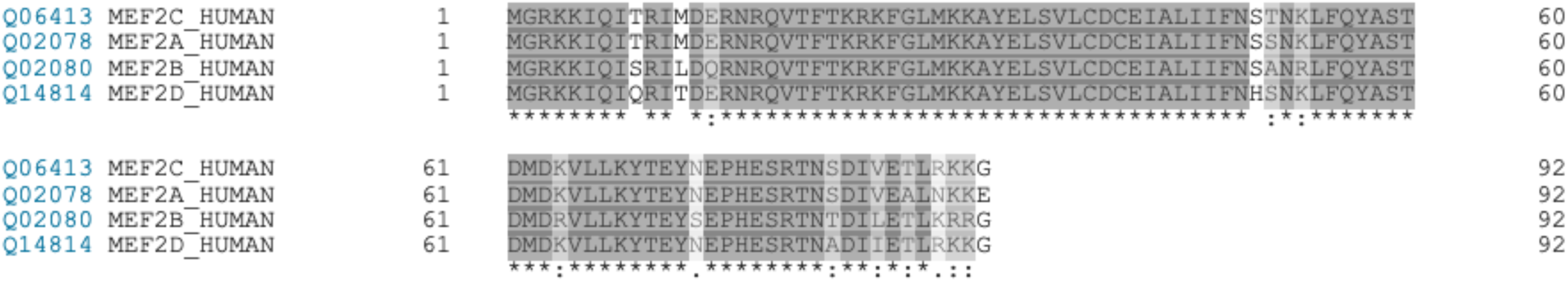
Protein sequence alignment of the four human myocyte enhancer factor 2 proteins proteins (MEF2A-D) using the Clustal-Omega default alignment function in UniProt for the first 92 N-terminal residues. Coloured by similarity; * = identical amino acids in all 4 proteins; : = similar amino acids in all 4 proteins.

**Supplementary Figure 5:**
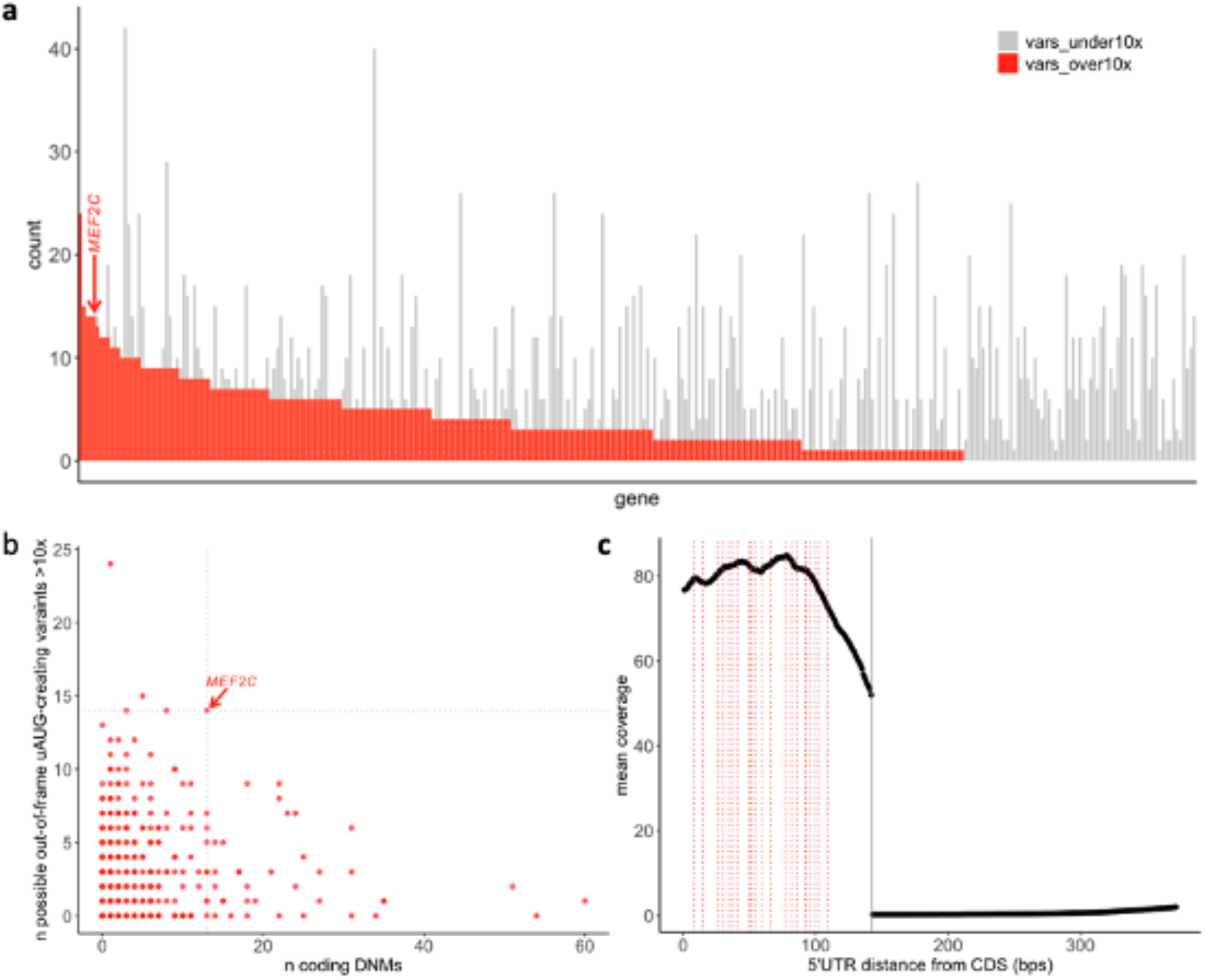
**Coverage of uAUG-creating sites of DD haploinsufficient genes and the *MEF2C* 5’UTR**. (a) Stacked bar chart showing the count of all possible uAUG-creating variants that would create out-of-frame overlapping ORFs that are covered at mean >10x (red), or ≤10x (grey) per gene. *MEF2C* has a high number of possible variants (n=14), all of which are well covered. (b) The number of well-covered uAUG-creating variants that would create out-of-frame overlapping ORFs plotted against the number of coding missense and protein-truncating *de novo* mutations (DNMs) per gene. *MEF2C* has both a high number of well-covered sites and a high diagnostic yield. (c) The mean coverage across the *MEF2C* 5’UTR. All possible uAUG-creating variants that would create either out-of-frame overlapping ORFs or CDS-elongations are plotted as dotted lines. The 5’UTR exon that is adjacent to the CDS is very well covered (mean >50x). NB: (a) and (b) do not include CDS-elongating variants as these would not be predicted to cause loss-of-function unless there is an important N-terminal structure or functional domain.

## Supplementary Tables

**Supplementary Table 1**: List of haploinsufficient developmental disorder genes and their MANEv0.91 transcripts used for analysis.

**Supplementary Table 2**: List of missense variants identified in DD cases. ClinVar variants are filtered to only those identified as de novo or with experimental evidence. Protein changes are with respect to the Ensembl canonical transcript ENST00000340208.5.

**Supplementary Table 3**: List of gnomAD v2.1.1 missense variants in MEF2 genes used from protein modelling. Protein changes are with respect to the Ensembl canonical transcript ENST00000340208.5.

**Supplementary Table 4**: Residues in the structure of MEF2A and their direction with respect to the bound DNA.

**Supplementary Table 5**: Comparing the proportion of DD and gnomAD variants that are in contact/pointing towards DNA to those that are distal or pointing away from the DNA-binding interface.

**Supplementary Table 6**: Change in Gibbs free energy (ΔΔG) of protein-DNA interaction and complex stability associated with missense variants in MEF2C.

## Genomics England Research Consortium

John C. Ambrose^1^; Prabhu Arumugam^1^; Emma L. Baple^1^; Marta Bleda^1^; Freya Boardman-Pretty^1,2^; Jeanne M. Boissiere^1^; Christopher R. Boustred^1^; Helen Brittain^1^; Mark J. Caulfield^1,2^; Georgia C. Chan^1^; Clare E. H. Craig^1^; Louise C. Daugherty^1^; Anna de Burca^1^; Andrew Devereau^1^; Greg Elgar^1,2^; Rebecca E. Foulger^1^; Tom Fowler^1^; Pedro Furió-Tarí^1^; Adam Giess^1^; Joanne M. Hackett^1^; Dina Halai^1^; Angela Hamblin^1^; Shirley Henderson^1,2^; James E. Holman^1^; Tim J. P. Hubbard^1^; Kristina ibáñez^1,2^; Rob Jackson^1^; Louise J. Jones^1,2^; Dalia Kasperaviciute^1^; Melis Kayikci^1^; Athanasios Kousathanas^1^; Lea Lahnstein^1^; Kay Lawson^1^; Sarah E. A. Leigh^1^; Ivonne U. S. Leong^1^; Javier F. Lopez^1^; Fiona Maleady-Crowe^1^; Joanne Mason^1^; Ellen M. McDonagh^1,2^; Loukas Moutsianas^1,2^; Michael Mueller^1,2^; Nirupa Murugaesu^1^; Anna C. Need^1,2^; Peter O’Donovan^1^; Chris A. Odhams^1^; Andrea Orioli^1^; Christine Patch^1,2^; Mariana Buongermino Pereira^1^; Daniel Perez-Gil^1^; Dimitris Polychronopoulos^1^; John Pullinger^1^; Tahrima Rahim^1^; Augusto Rendon^1^; Pablo Riesgo-Ferreiro^1^; Tim Rogers^1^; Mina Ryten^1^; Kevin Savage^1^; Kushmita Sawant^1^; Richard H. Scott^1^; Afshan Siddiq^1^; Alexander Sieghart^1^; Damian Smedley^1,2^; Katherine R. Smith^1,2^; Samuel C. Smith^1^; Alona Sosinsky^1,2^; William Spooner^1^; Helen E. Stevens^1^; Alexander Stuckey^1^; Razvan Sultana^1^; Mélanie Tanguy^1^; Ellen R. A. Thomas^1,2^; Simon R. Thompson^1^; Carolyn Tregidgo^1^; Arianna Tucci^1,2^; Emma Walsh^1^; Sarah A. Watters^1^; Matthew J. Welland^1^; Eleanor Williams^1^; Katarzyna Witkowska^1,2^; Suzanne M. Wood^1,2^; Magdalena Zarowiecki^1^.

^1^Genomics England, London, UK

^2^William Harvey Research Institute, Queen Mary University of London, London, EC1M 6BQ, UK.

